# Predicting COVID-19 Vaccine Efficacy from Neutralizing Antibody Levels

**DOI:** 10.1101/2021.10.13.21264921

**Authors:** Majid R. Abedi, Samuel Dixon, Timothy Guyon, Serene Hsu, Aviva R. Jacobs, Lakshmi Nair, Robert Terbrueggen

## Abstract

Recent studies using data accrued from global SARS-CoV-2 vaccination efforts have demonstrated that breakthrough infections are correlated with levels of neutralizing antibodies. The decrease in neutralizing antibody titers of vaccinated individuals over time, combined with the emergence of more infectious variants of concern has resulted in waning vaccine efficacy against infection and a rise in breakthrough infections. Here we use a combination of neutralizing antibody measurements determined by a high throughput surrogate viral neutralization test (sVNT) together with published data from vaccine clinical trials and comparative plaque reduction neutralization test (PRNT) between SARS-CoV-2 variants to develop a model for vaccine efficacy (VE) against symptomatic infection. Vaccine efficacy estimates using this model show good concordance with real world data from the US and Israel. Our work demonstrates that appropriately calibrated neutralizing antibody measurements determined by high throughput sVNT can be used to provide a semi-quantitative estimate of protection against infection. Given the highly variable antibody levels among the vaccinated population, this model may be of use in identification of individuals with an elevated risk of breakthrough infections.

## Introduction

While Pfizer and Moderna vaccine protection against hospitalization and severe disease remains high at ∼90%, an increasing number of fully vaccinated individuals are experiencing breakthrough COVID-19 infections. The August SARS-CoV-2 Infections and Hospitalizations reports (May 1-July 25, 2021) for New York City^1^ and Los Angeles County^2^ showed a combined total of 20,570 breakthrough cases (>20% of total infections for NY and LA during this period), compared with only 10,262 total breakthrough cases^3^ for the entire United States from January 1-April 30, 2021 (<0.2%). The recent CDC HEROES-RECOVER report^4^ of frontline health workers showed a vaccine efficacy against infection by the Delta variant of 66% compared to 91% in the months preceding Delta dominance, and Israel reported that VE against symptomatic Delta variant infection for fully vaccinated Pfizer subjects decreased to 57% after 6 months^5^. Israel began offering booster shots at the beginning of August and medical officials recently reported that a third dose increased NAb more than 10-fold^6^, vaccine efficacy against infection to 86%^7^ and resulted in a 11.4-fold decrease in relative risk of infection compared to two doses only^8^.

There is growing evidence that the decrease in vaccine efficacy against symptomatic infection is being driven by both a decrease in neutralization efficacy by vaccine antibodies against the Delta variant^9^ and waning NAb titer^10^. Current data suggest that NAb titer at time of exposure is a primary determinant for vaccine efficacy against infection^11^, while protection against hospitalization/severe disease is more dependent on an individual’s ability to make new reinforcing neutralizing antibodies via memory B cells and T cell mediated immune protection in the days following infection. Khoudry^11^ et al has previously developed a model for predicting vaccine efficacy based on neutralizing antibody levels using data from 7 phase 3 COVID-19 vaccine clinical trials, and they subsequently proposed using the difference in PRNT fold-dilution between wild type (WT) and variant SARS-CoV-2 strains as a “correction factor” to account for different strains^12^. Alpha (B.1.1.7, UK variant) has a best fit correction factor of 1.6, Beta (B.1.351, South African Variant) a correction factor of 8.8, and Delta (B.1.617.2) of 3.9.

Here we apply the approach outlined above to develop a model for predicting vaccine efficacy using NAb titer and decay kinetics obtained from an EUA authorized high throughput Blocking Enzyme-Linked Immunosorbent Assay (ELISA) for a cohort of Pfizer and Moderna vaccinated subjects from 0 to 250 days post-full vaccination. We then test the model against real world studies of observed vaccine efficacy for preventing symptomatic infection for Moderna and Pfizer immunized subjects

## Results

### Development of Neutralizing Antibody Titer versus Vaccine Efficacy Model

A cohort of 582 study participants including 331 fully vaccinated Pfizer, 174 fully vaccinated Moderna, 49 prior positive (convalescent), and 28 J&J subjects were included in the study with times post-full-vaccination spanning 0 to 250 days. Venous draw (EDTA) and matched fingerstick (DxCollect MCD Serology) specimens were collected under IRB and the plasma was tested for wild-type (WT) SARS-CoV-2 neutralizing antibodies (NAbs) using a high-throughput blocking ELISA, surrogate viral neutralization test (sVNT)^13^.

As described previously^11^, a percent vaccine efficacy (%VE) model was developed based on the measured mean NAb levels for Pfizer, Moderna and J&J vaccinated subjects during first 30 days post-full vaccination and the reported Phase 3 clinical trial efficacy against symptomatic infection (Figure 1).

**Figure 1:**
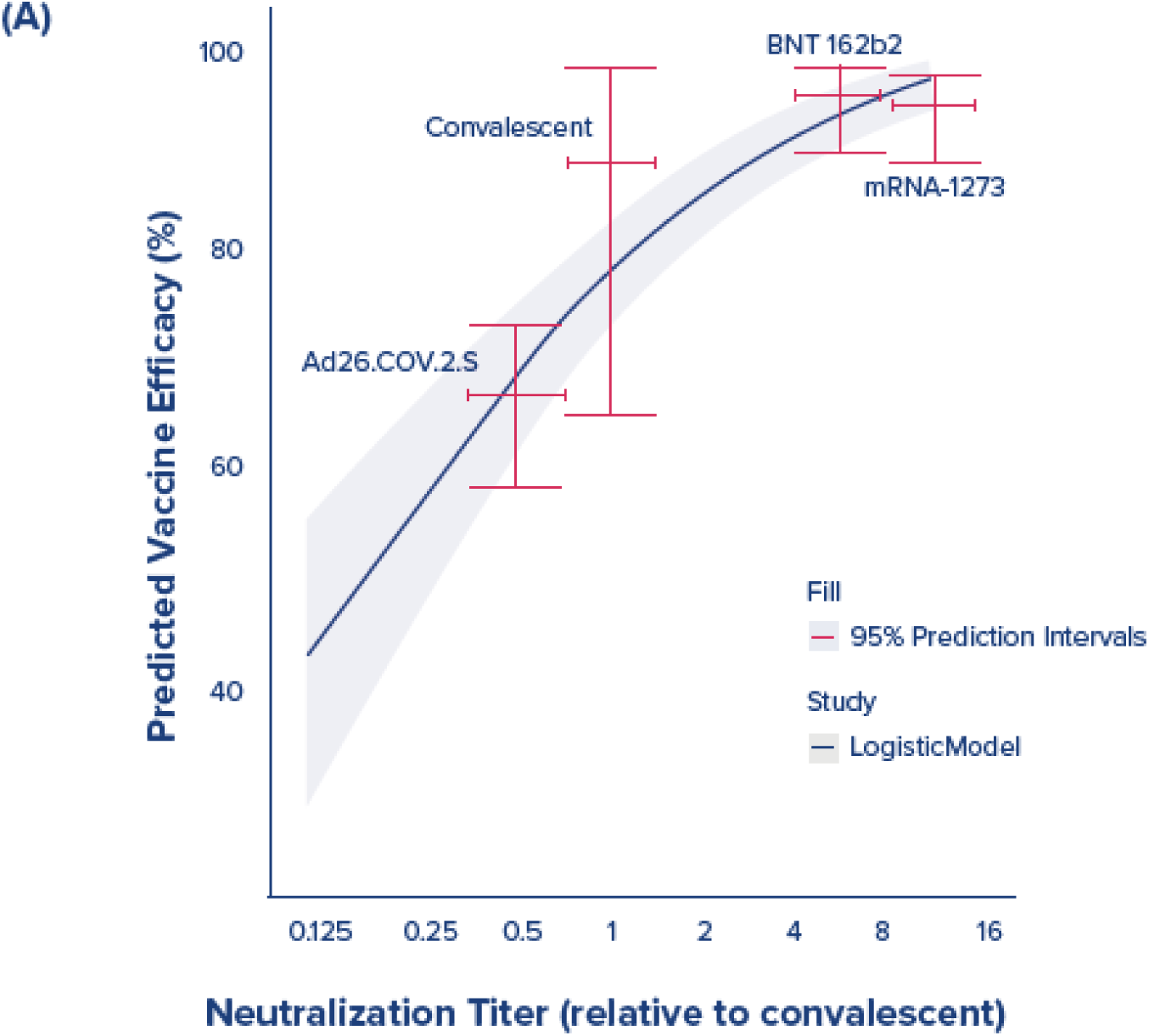
Vaccine Efficacy Against Symptomatic Infection is based on the logistic model developed by Khoury et al using calibration to mean NAb titer measured for Moderna, Pfizer, J&J and convalescent (former positive) subjects.

### Correlation of Neutralizing Antibodies to Plaque Reduction Neutralization Test (PRNT) assay

A sub-cohort of 132 plasma samples from vaccinated and convalescent patients was analyzed by PRNT. Probit analysis was performed on data to calculate PRNT50 (IC50) values (Table 1). A line was fit to the log transformed data (Figure 2A) and good correlation (R^2^=0.68) was observed for NAb levels for all samples regardless of whether the NAbs were produced by infection or vaccination. The fit improved (R^2^=0.82) when restricted to the 75 vaccinated samples (Figure 2B), most likely due to the increased uniformity of vaccine generated antibodies. Figure 2C shows a plot of log transformed mean NAb titer versus mean of Probit IC50 by vaccine and prior infection with an excellent correlation (R^2^ = 0.95). Taken together, the data illustrates that NAb titers as measured by sVNT are highly correlated with virus neutralizing effect by PRNT.

**Table 1:** Summary table of number of samples, mean NAb titer ± standard deviation, and mean PRNT Probit IC50 ± standard deviation for samples sent for PRNT testing.

| Vaccine Manufacturer | # Samples | Mean(NAb Titer) | Mean(Probit Estimated IC50) |
| --- | --- | --- | --- |
| Moderna | 21 | 13404 (± 10348) | 964 (± 1004) |
| Pfizer | 34 | 6130 (± 5355) | 401 (± 454) |
| J&J | 28 | 916 (± 740) | 50 (± 46) |
| Convalescent | 49 | 2132 (± 1449) | 230 (± 167) |

**Figure 2:**
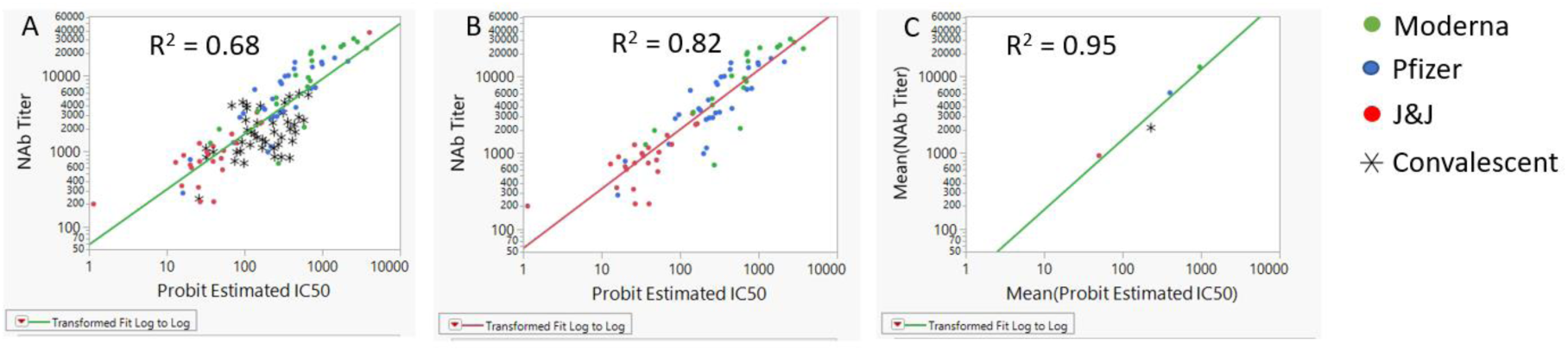
Log-Log plots of neutralizing antibody titer versus Probit IC 50. (A) all 132 samples analyzed with PRNT, (B) Post-vaccination samples only, (C) Mean titer by infection and vaccine type

### Decline of Moderna and Pfizer neutralizing antibody titer with time post-vaccination

The NAb titers of Moderna and Pfizer vaccinated subjects were analyzed to determine the drop in NAb titers as a function of time post-full vaccination. Only fully vaccinated subjects with no indication of prior infection were included. In order to test for evidence of prior infection, a subset of samples was tested for IgG antibodies for the Nucleoocapsid (N) protein, and samples suspected of being from previously infected donors were excluded. A total of 174 Moderna subjects and 331 Pfizer subjects through 7 months post-vaccination were included (Table 2). Mean NAb titers for Moderna are consistently approximately 2-fold higher than mean NAb titers from Pfizer vaccinations. There is a significant decline in average antibodies over time and the data also shows a significant amount of variability (Figure 3). NAb decay for Moderna and Pfizer were modeled using first order decay kinetics yielding similar half-lives of 74.5 and 71.7 days, respectively. The combined half-life value of 72.3 days [66.5-80.4] was used to develop models for predicting average NAb titer as a function of days post-full vaccination.

**Table 2:** Number of Moderna and Pfizer fully vaccinated subjects by month post-full vaccination. Full vaccination is based on 14 days after the 2^nd^ dose.

|  | # Moderna Vaccinated | Moderna | # Pfizer Vaccinated | Pfizer |
| --- | --- | --- | --- | --- |
| Month 0 | 37 | 24214 | 35 | 12090 |
| Month 1 | 14 | 18376 | 67 | 11273 |
| Month 2 | 29 | 17744 | 35 | 8671 |
| Month 3 | 25 | 13218 | 51 | 7640 |
| Month 4 | 17 | 9825 | 64 | 4331 |
| Month 5 | 28 | 5347 | 19 | 3312 |
| Month 6 | 23 | 6342 | 25 | 2128 |
| Month 7 |  |  | 33 | 2116 |

**Figure 3:**
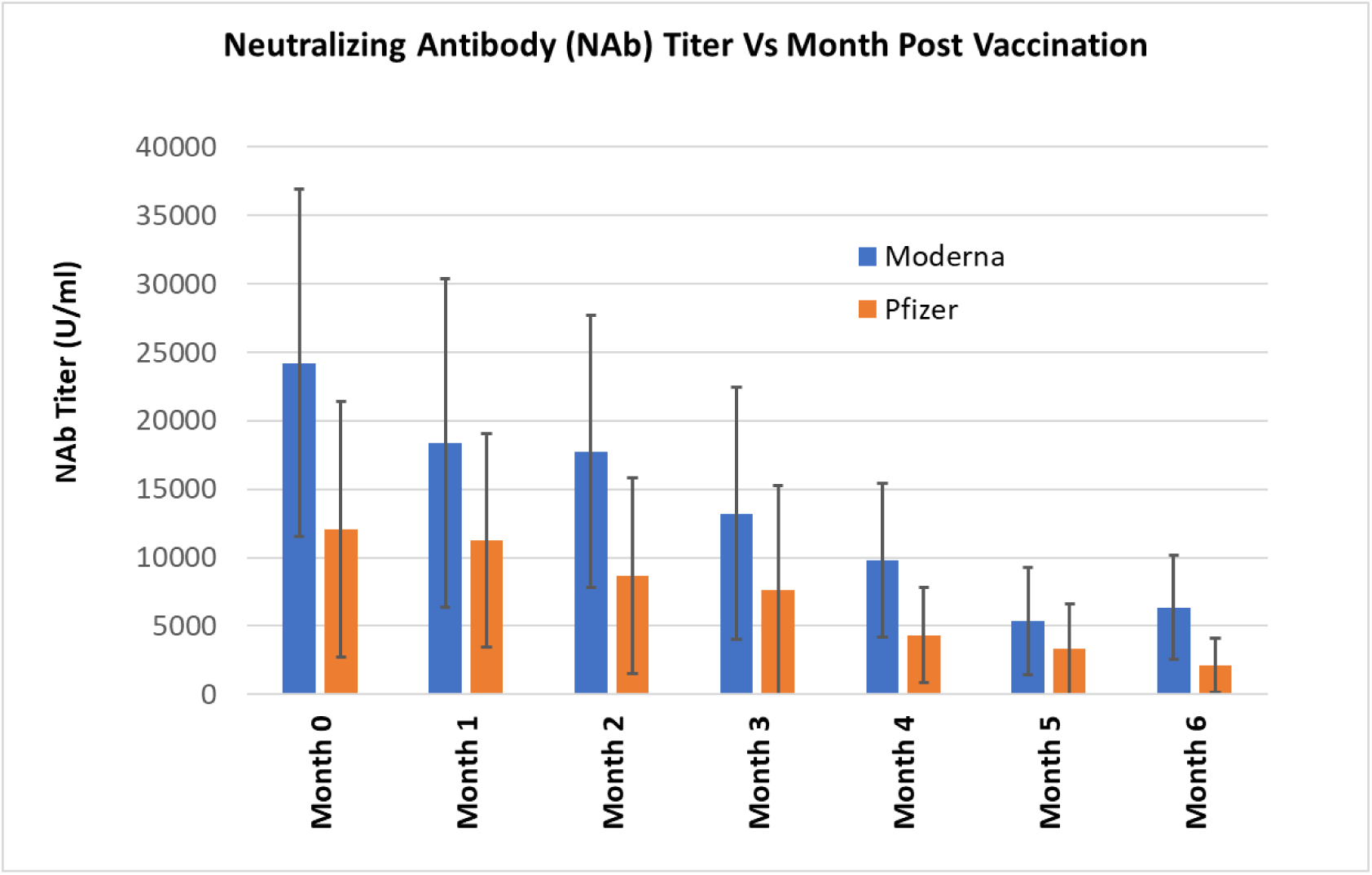
Plot of sVNT neutralizing antibody titer with error bars for Moderna and Pfizer vaccinated subjects.

### Immunity Assessment Test (IAT) VE Decay Model

We developed an Immunity Assessment Test (IAT) VE Decay Model to estimate vaccine efficacy over time (see Material and Methods for details). To adjust for the difference in VE between the WT type strain used in the initial VE model and the Delta variant, we divided the estimated NAb titer by 3.9^12^, which is based on the fold difference in Plaque Reduction Neutralization Test (PRNT) observed for the same NAb containing sample when exposed to Wild Type and Delta Variant SARS-CoV-2. Figure 4 shows the average estimated VE for the Delta Variant as a function of months post-full vaccination. The IAT VE Decay Model predicts that initial vaccine efficacy for Moderna is 90% and Pfizer is 85% against the Delta variant immediately following full vaccination. At month 6 post-full vaccination, VE against Delta is predicted to decrease to 62% for Pfizer and 75% for Moderna.

**Figure 4:**
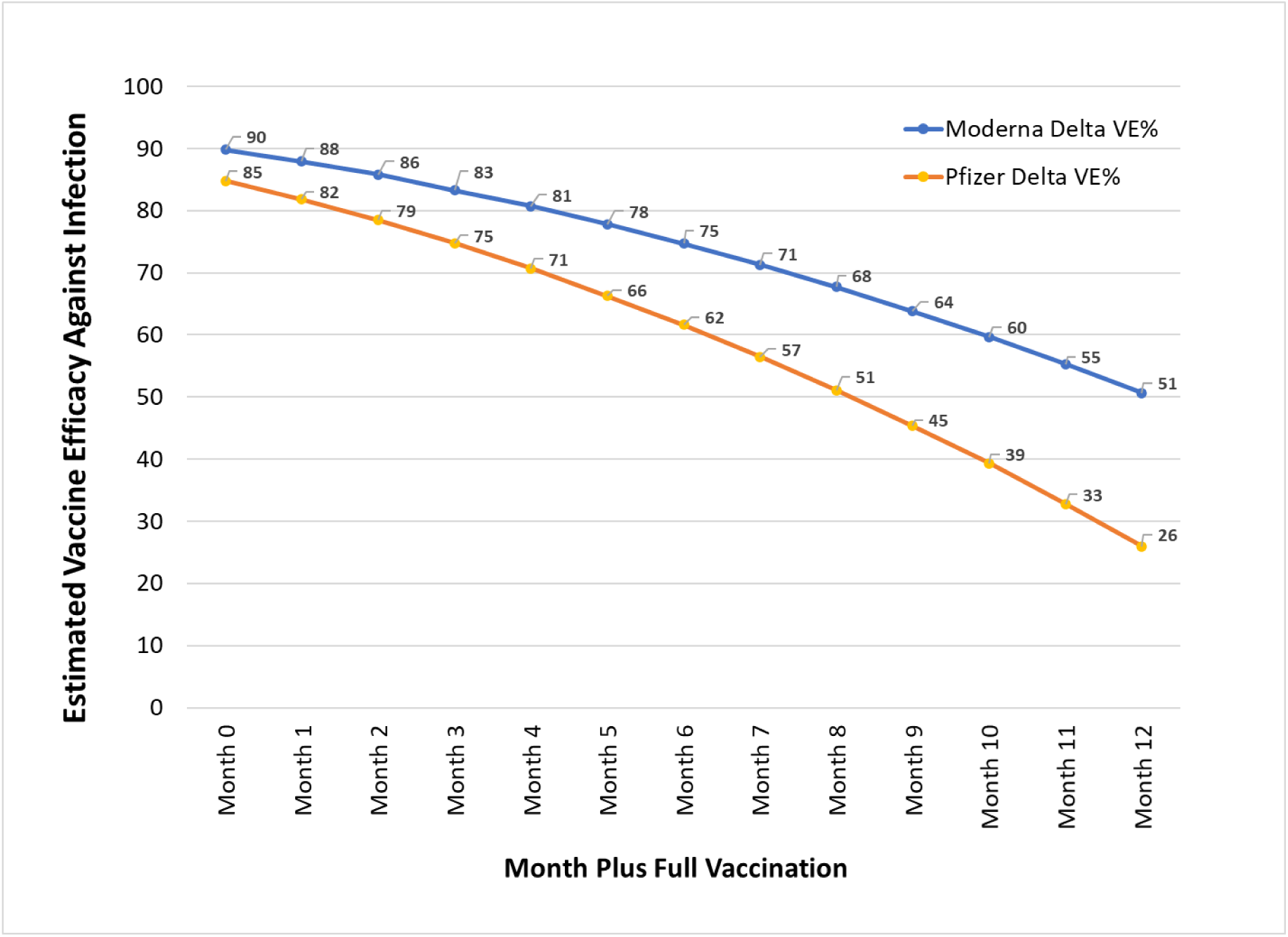
IAT VE Decay Model Estimated vaccine efficacy (VE) against symptomatic infection from the Delta variants of SARS-CoV-2 versus months post-full vaccination for Moderna and Pfizer vaccines.

### Comparison to Real World Data

In order to evaluate the accuracy of the IAT VE Decay Model, the predicted VE values were compared to the reported VE from large population studies reported in the literature. Israel was the earliest country to vaccinate more than 50% of its population, reaching this level by March of 2021 compared to June or later for other developed countries. The COVID-19 incidence rate dropped from 900 cases per million per day in mid-January 2021 to less than 2 cases per million per day by June 2021^14^. The country re-opened completely and was well on their way to normalcy when the Delta Variant emerged and became the dominant strain by mid-June. Israel reported^15^ a drop in Pfizer vaccine efficacy to 64% on July 5^th^ and recently published a study showing a decrease in Pfizer vaccine efficacy to 57% against infection after 6 months post-vaccination^16^. Figure 5 shows a plot of predicted Pfizer VE versus reported VE from Israel^16^ aboveobserved for months 2-6 months post-vaccination with 95% confidence levels. The real world VE values reported from Israel and predicted %VE values from the IAT VE Decay model are highly concordant with the reported VE values all falling within the 95% CI of the estimated values.

**Figure 5.**
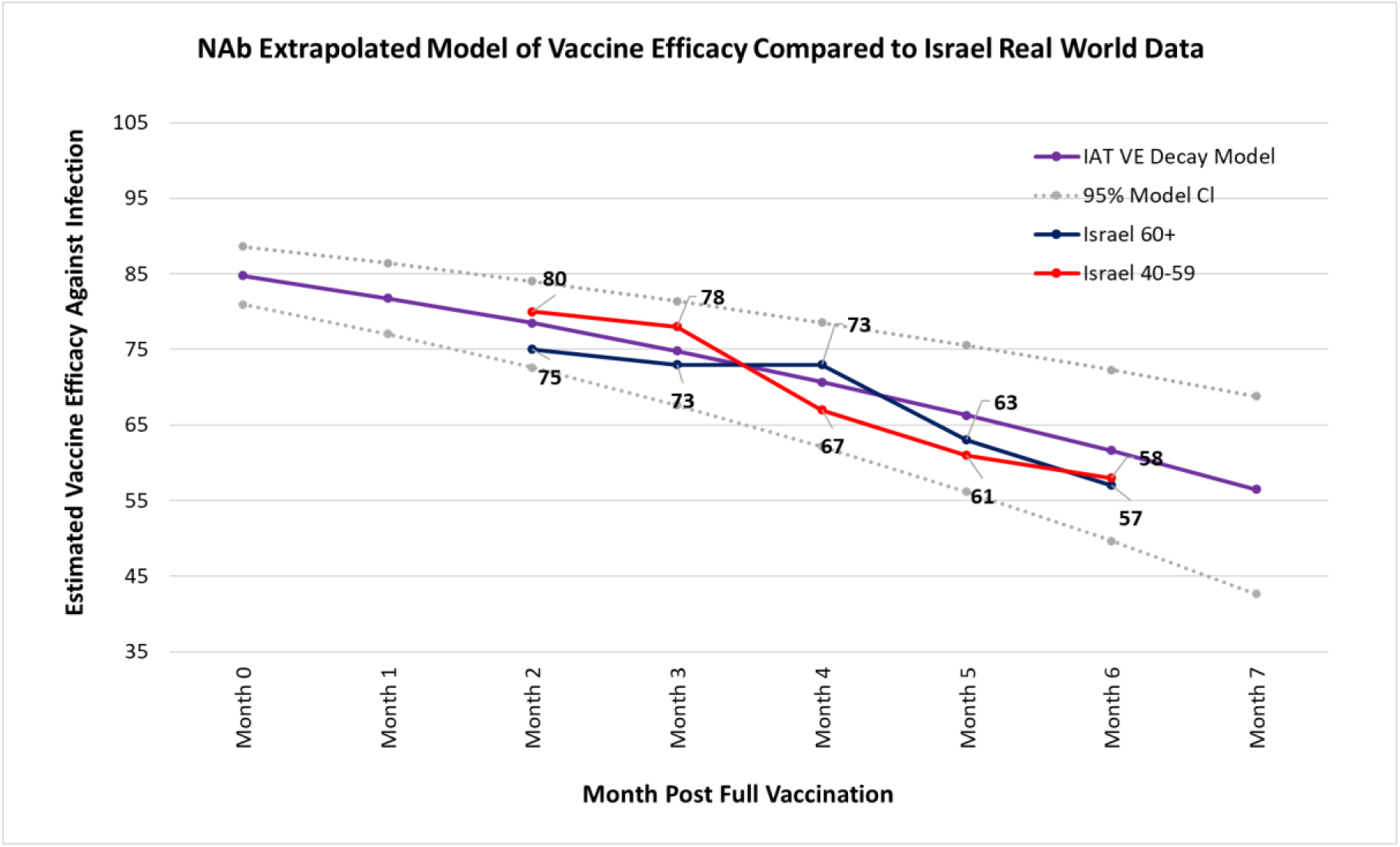
Estimated VE from the IAT VE Decay Model assuming Pfizer vaccination and protection form WT SARS-CoV-2 compared to reported VE from Israel. Dashed lines show 95% CI for the IAT VE Decay Model values.

#### Los Angeles Real World Cohort

The Los Angeles County Real World Data Cohort^17^ provides vaccine manufacturer specific numbers for breakthrough infections and the average vaccination prevalence by vaccine. LA county was 56.5% vaccinated of which 53% was Pfizer, 40% Moderna and 7% J&J. Of the 10,895 breakthrough infections between May 1 and July 25^th^, 16.8% were in J&J – 2.4 times J&J prevalence. Using an estimated LA county population of 9.95 million, we calculate an uncorrected VE of 81% Moderna, 71.6% Pfizer and 34.7% J&J.

During the period of May to July 2021, the distribution of VOC changed from mostly Alpha to >90% Delta variant. In order to generate estimated VE using our model, we factored in both the time post-vaccination and also the changing prevalence of Delta and Alpha variants (together with the corresponding differences in VE for the Alpha and Delta variants). The estimated VE was 79.9% for Moderna and 68.3% for Pfizer which are concordant with the real-world values (shown in Table 3).

**Table 3.** IAT VE Decay Model Estimated VE and Reported Vaccine Efficacy measurements (May 1–July 25) from Los Angeles County^17^

| Vaccine | Measured VE% | Estimated VE% |
| --- | --- | --- |
| Moderna | 81.0% | 79.9% |
| Pfizer | 71.6% | 68.3% |
| J&J | 34.7% | Insufficient Data Available |

#### CDC Heroes Recover Cohort

The CDC HEROES-RECOVER report^18^ of frontline health workers showed a vaccine efficacy against infection by the Delta variant of 66% compared to 91% in the months preceding Delta dominance. The Delta predominance period is estimated from June 26, 2021 to August 14, 2021 using CDC variant data with an average predominance of 85.2% Delta and 14.8% Alpha. The cohort was 65% Pfizer, 33% Moderna and 2% J&J. Considering the relative proportions of VOCs as well as the type of vaccines, the IAT VE Decay Model estimated a VE of 65.5% versus the real world reported value of 66% (Table 4).

**Table 4.** IAT VE Decay Model Estimated VE and Measured Vaccine Efficacy Measurements (Jun 26 – Aug 14) from the CDC Heroes Recover Cohort^18^

| Vaccine | Measured VE% | Estimated VE% |
| --- | --- | --- |
| Pfizer/Moderna/J&J<br>(65%/33%/2%) | 66.0% | 65.5% |

## Discussion

Results from our study demonstrate that neutralizing antibodies produced by either vaccination or infection show a good correlation with PRNT testing. Our data also shows a clear trend that single dose J&J vaccinated individuals produce substantially less NAbs compared to fully vaccinated Pfizer and Moderna subjects which would be expected to result in lower vaccine efficacy for SARS-CoV-2 infection. These results are consistent with Phase 3 clinical trial results as well as a recent report from the CDC^19^ that demonstrated 32.7% breakthrough infections for fully vaccinated J&J participants, compared to just 17.3% and 11.3% for fully vaccinated Pfizer and Moderna, respectively. The low level of starting NAbs for J&J vaccinated individuals is concerning, especially considering the ∼4-fold reduced neutralizing efficacy of vaccine antibodies for the Delta variant compared to WT COVID-19. J&J vaccinated individuals showed an uncorrected vaccine efficacy of only 34.7% in the recent LA data^17^, suggesting that a booster dose may be warranted immediately.

For Moderna and Pfizer vaccinated individuals, there is a clear trend of decreasing NAbs with time. The 72.3 [95% Cl 66.5-80.4] day NAb half-life for Moderna and Pfizer is consistent with studies reporting between 65 days for mRNA vaccine induced NAbs and 108 days for infection produced antibodies^20,21^. The high standard deviation of the measured NAb titers (Figure 3) suggest significant differences in levels of NAb response to vaccination and highlights the fact that the current policy of recommending booster shots after 6-months post vaccination for Pfizer will almost certainly result in people who have lowered VE not receiving a 3^rd^ dose.

Our data also shows that post-vaccination, NAb titers are approximately 2-fold higher for Moderna as compared to Pfizer. This, in part, contributes to the increased vaccine efficacy observed for Moderna versus Pfizer vaccines. Given the similar mechanisms of action between the 2 mRNA vaccines, it is possible that the increased starting titers may be related to the higher Moderna vaccine dose (100ng compared to 30ng for Pfizer) as well as the difference between the 1^st^ and 2^nd^ dose (4 weeks vs 3 weeks). This may have implications on the recent reports that the FDA is considering approving Moderna booster shots at 50ng^22^.

The IAT VE Decay Model was generated using NAb titers obtained from convalescent and fully vaccinated participants using a high throughput semi-automated sVNT workflow. The resulting VE estimates over time (Figure 4, and Tables 3 & 4) are remarkably consistent with the real-world data that has been published by Israel^16^ as well as real world studies published by the CDC^18^ and the county of Los Angeles^17^. This demonstrates the utility of sVNT generated NAb titers to calculate VE and has the added advantage over virology-based tests (such as PRNT and Pseudovirus Neutralization Assays) in that it is amenable to high throughput automation and the time to results are short. While the IAT VE Decay Model may require refinement as more real-world VE data becomes available, it can be considered as a useful tool in assessing immune protection from NAb titers obtained from individual participants.

Taken together, our data highlight the benefits of using NAb titers to assess vaccine efficacy and in addition supports the conclusion that after 6-8 months post-vaccination, booster doses are warranted Pfizer, while Moderna vaccinated individuals appear to have a higher level of protection.

## Conclusion

The potential impact of emerging variants on VE is estimated with reasonable accuracy by comparative testing of wild type and emerging variant strains using PRNT. Predictive models can be subsequently adjusted by dividing the NAb titer variable by the fold dilution impact as measured by PRNT IC50.

NAbs for Pfizer and Moderna vaccines show similar rates of decay, and Moderna’s observed increased VE relative to Pfizer appears to be primarily due to the higher (∼2x) initial NAb levels produced following full vaccination, and possibly by a slightly slower decay rate. Pfizer booster doses after month 6 seems reasonable, while Moderna vaccinated individuals may not require boosting until 1 year or longer if the vaccine cocktail is adjusted to account for new variants.

The close agreement between vaccine efficacy estimated by the NAb based VE model and real-world data strongly supports the conclusion that NAb titer is a strong correlate of protection against symptomatic infection. While current available VE data allows for general guidelines on fixed-interval post-vaccination booster shots to be proposed, variations in individual vaccine response and uncertainty in decay kinetics suggest a need for individualized testing.

## Materials and Methods

All samples were collected into DxTerity DxCollect MCD Serology tubes following finger puncture or K2EDTA using standard phlebotomy techniques. All subjects were consented under clinical research protocols reviewed and approved by an accredited Institutional Review Board (IRB) and implemented in accordance with the ICH Harmonized Guidelines for Good Clinical Practice (GCP), applicable regulations (including CFR Title 21). Subject enrollment occurred at various clinical sites in Arizona and Southern California. De-identified clinical information including demographics, previous COVID infection and vaccination status was collected.

### Plasma Isolation

Plasma was isolated by spinning the K2EDTA tubes in a benchtop centrifuge at 2,000 rpm for 10 minutes and aspirating the top-most liquid layer which was transferred to a 5ml cryo-storage vial. For MCD Serology fingerstick blood collection devices, the stabilized blood sample was aspirated from the device into a 1.5mL sterile microfuge tube and centrifuged at 3,500 rpm for 5 minutes. The plasma supernatant was then transferred to a fresh 1.5 ml microfuge tube.

### Neutralizing antibody testing

Neutralizing antibody testing was performed using a semi-automated workflow with 96-well plates and liquid handling stations. Sample input for the Genscript cPass (GenScript P/N A02087) kit was generated by diluting the K2EDTA plasma 10-fold (6μL K2EDTA plasma combined with 54μL Sample Dilution Buffer). For MCD Serology fingerstick blood samples, the MCD Serology plasma was diluted to match the total protein concentration of 10-fold diluted K2EDTA Blood (equivalent to 6.5mg/mL). Briefly, MCD Serology Plasma samples were tested with the Pierce BCA Protein Assay Kit (Thermo Scientific, P/N 23225) to determine the total protein content and the appropriate volume of plasma and sample dilution buffer was added to result in a final concentration of 6.5mg/mL total protein in 60μL.

Neutralization Antibody Testing was conducted following the Instructions for Use of the EUA authorized “cPass SARS-CoV-2 Neutralization Antibody Detection Kit” with the exception that a neutralizing antibody standard curve generated from the GenScript SARS-CoV-2 Neutralizing Antibody Standard (GenScript P/N A02087) was included on the plate to allow for quantitative determination of neutralizing antibody titers. Briefly, the diluted plasma sample was incubated at 37°C for 30-minutes with a solution of HRP-RBD. The Plasma/HRP-RBD solution was then transferred to a capture plate coated with anti-RBD antibodies and incubated at 37°C for 15-minutes. The capture plate was then washed 3-times with Wash Buffer and TBM solution (HRP substrate) added to each well for 15-minutes followed by addition of Stop Solution and detection at OD450. Neutralizing antibody standards of known titers; a Positive Control containing anti-RBD neutralizing antibodies; and a Negative control were included in duplicate in every run. Neutralizing antibody titers were determined by comparison to the standard curve. In cases where the OD450 value fell outside the upper limit of quantification of the standard curve, the sample was diluted 6-fold in Sample Dilution Buffer (provided in the cPass Kit) and re-tested.

### IAT VE Decay Model Development

#### Vaccine Effectiveness Model

A predictive model of immune protection from COVID-19 was generated using the methodology described by Khoury et al^20^. Briefly, Neutralizing antibody titers from vaccinated individuals were normalized to the results of convalescent COVID-19 patients and vaccine efficacy results observed in phase III clinical trials of three vaccines (mRNA-1273, BNT162b2 and Ad26.COV2.S) were fit using a logistic model. Analysis was performed using R code adapted from code published by the Khoury group^23^.

#### Neutralizing Antibody Decay

A non-linear, single order exponential decay model was fit to population data of neutralizing antibody titer vs days post-full vaccination using JMP 15^24^ and the half-life was calculated for two vaccines (mRNA-1273 and BNT162b2).

#### Adjustment for Delta Variant

An adjustment to compensate for the antigenic difference between the wild type antigen represented in each vaccine and the currently circulating delta variant is necessary. We compensated for this difference using the methodology described by Cromer et al.^12^ Briefly, the mean Neutralizing antibody titers were calculated by the single order exponential decay model for each vaccine up to 8 months post vaccination. Mean neutralization titers were adjusted by reported decrease in neutralizing antibody effectiveness for the delta variant. The resulting adjusted titer was then input into the vaccine effectiveness model to calculate the estimated vaccine effectiveness at a given time post vaccination. Mean results for two vaccines were then compared to real-world vaccine effectiveness observations in the United States and Israel.

## Data Availability

All data produced in the present study will be made available at reasonable request to authors once published in a peer reviewed journal

